# Severe Acute Respiratory Syndrome Coronavirus 2 detection in induced sputum of asthmatic patients using saliva sampling device

**DOI:** 10.1101/2022.12.12.22283341

**Authors:** Catherine Moermans, Sara Gerday, Noémie Bricmont, Romane Bonhiver, Florence Schleich, Julien Guiot, Makon Sébasien Njock, Monique Henket, Françoise Guissard, Virginie Paulus, Emmanuel Di Valentin, Frédéric Minner, Laurent Gillet, Fabrice Bureau, Renaud Louis

## Abstract

Since the beginning of the severe acute respiratory syndrome coronavirus 2 pandemic, the potential contamination of the induced sputum obtained from asthmatic patients in routine is a question of concern. The goal of this study was to assess this contamination using a saliva sample collection device. One hundred seventy-five sputum samples of asthmatic patients without fever were tested. We did not identify any positive PCR on sputum samples from asthmatic patients reporting chronic/episodic respiratory symptoms similar to what is seen in case of COVID-19. This technique was useful to evaluate the contamination of sputum samples generated during the pandemic.

## 1. Introduction

In the beginning of 2020, a new pandemic caused by Severe Acute Respiratory Syndrome Coronavirus 2 (SARS-CoV-2) inducing coronavirus infectious disease 19 (COVID-19) has spread around the world. This highly infectious virus was responsible for more than 5.5 millions of deaths around the world including more than 28,000 deaths in Belgium (World Health Organization, January 2022). The wide range of clinical manifestations of COVID-19 can vary from asymptomatic infection to severe pneumonia or acute respiratory distress syndrome. Common symptoms include cough and breathing difficulties usually with fever and fatigue. However, even asymptomatic individual can spread the virus, mainly by transmission that involve respiratory secretions.

Asthma is the most common chronic inflammatory respiratory disease worldwide [1]. The symptoms include wheeze, shortness of breath, chest tightness, cough, associated with reversible airflow limitation (http://ginasthma.org/). Regarding asthma and COVID-19 risk of infection, apart those receiving oral corticosteroids (OCS) as maintenance [2], asthma was not shown to be a risk factor for intensive care unit (ICU) admission and death related to COVID-19 [3] neither for hospitalization and ventilator use [4].

Induced sputum remains the gold standard to assess the airway inflammation in asthma, which is recommended in the management of severe asthma by the European Respiratory Society (ERS) and American Thoracic Society (ATS) guidelines [5]. Nasopharyngeal and throat swabs have been extensively used to detect SARS-CoV-2 in the population, however, sputum has been shown to contain a higher virus load that can be detected even after nasopharyngeal sample turned negative [6].

In addition, sputum seems more reliable and has a lower false-negative rate than upper respiratory sampling [7].

In our pneumology department, we have been using the non-invasive technique of induced sputum to monitor and phenotype the airway inflammation of the asthmatic patients since more than 20 years and we continued to collect induced sputum during the pandemic. We, however, adapted the induction of the sputum since February 2020. The induction procedure was first stopped and resumed in June in a new environment which was directly connected to outside air and the recommendations published in 2020 describing the sputum induction biosafety during the pandemic were followed [8]. However, the potential contamination of these samples from asthmatic patients presenting respiratory symptoms close to what is observed in case of symptomatic COVID-19 infection was a question of concern.

Samples coming from sputum induced during the pandemic, such as supernatants or cell pellets, are potentially contaminated and cannot be used for further experiments after Biosafety level (BSL)-2 processing. Indeed, those samples cannot be manipulated outside a BSL-2 or BSL-3 facility for research activities such as proteins or gene expression measurement (among other) using sputum supernatants or cells respectively. There is a need for an easy way to make sure that there are not suspect and therefore that their usage can be retrieved.

## 2. Material and methods

One hundred seventy-five sputum samples of asthmatic patients from our asthma clinic who underwent investigation as part of routine clinical practice, induced between November 2019 and October 2021, were randomly selected. Asthma was diagnosed in 157/175 patients according to the Global Initiative for Asthma guidelines (http://ginasthma.org/). None of the tested patients presented with overt acute respiratory infection or fever at time of sputum sampling. The absence of fever was checked using a forehead thermometer. Patient demographic and functional characteristics as well as respiratory symptoms are displayed in Table 1.

**Table 1.**
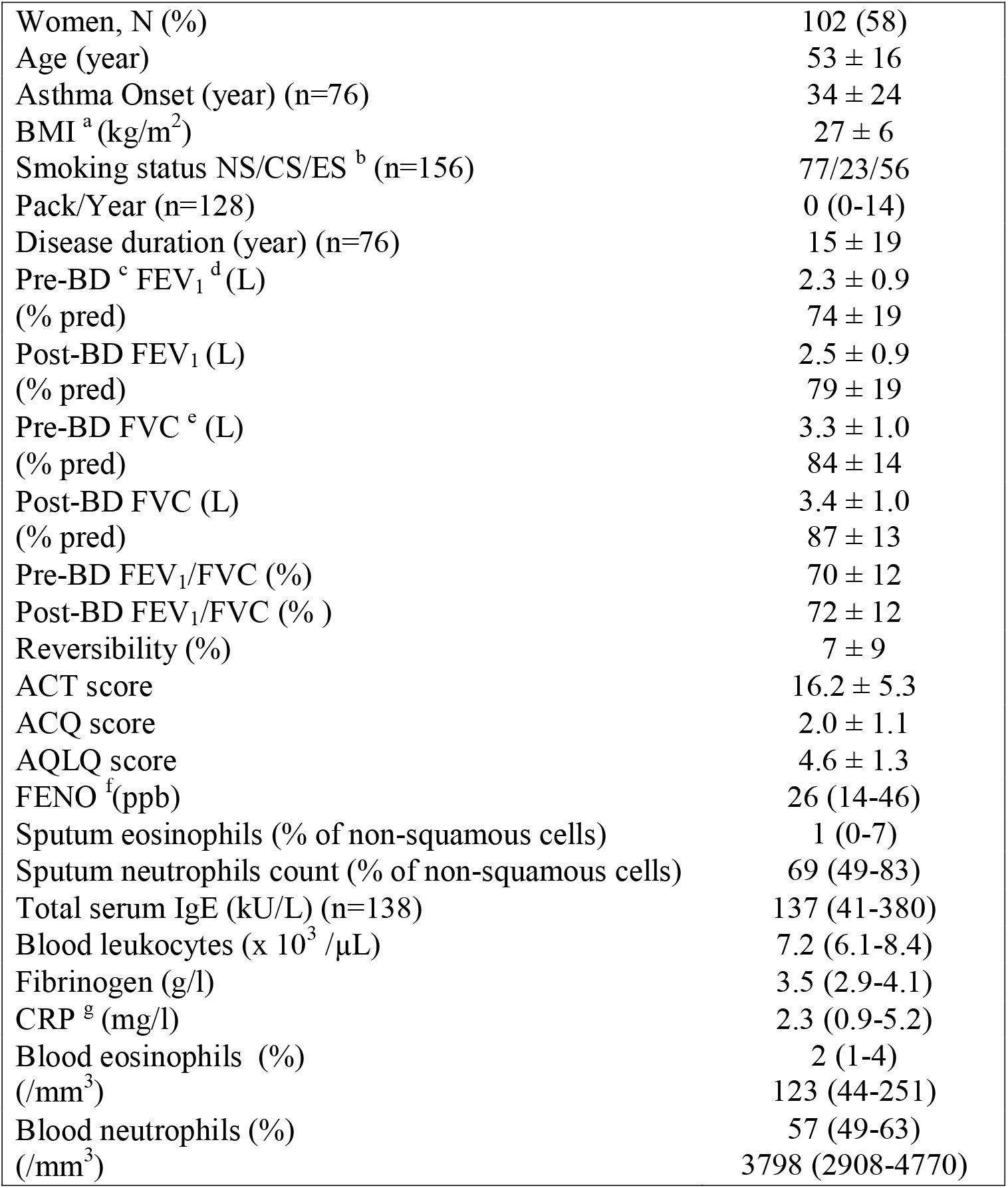
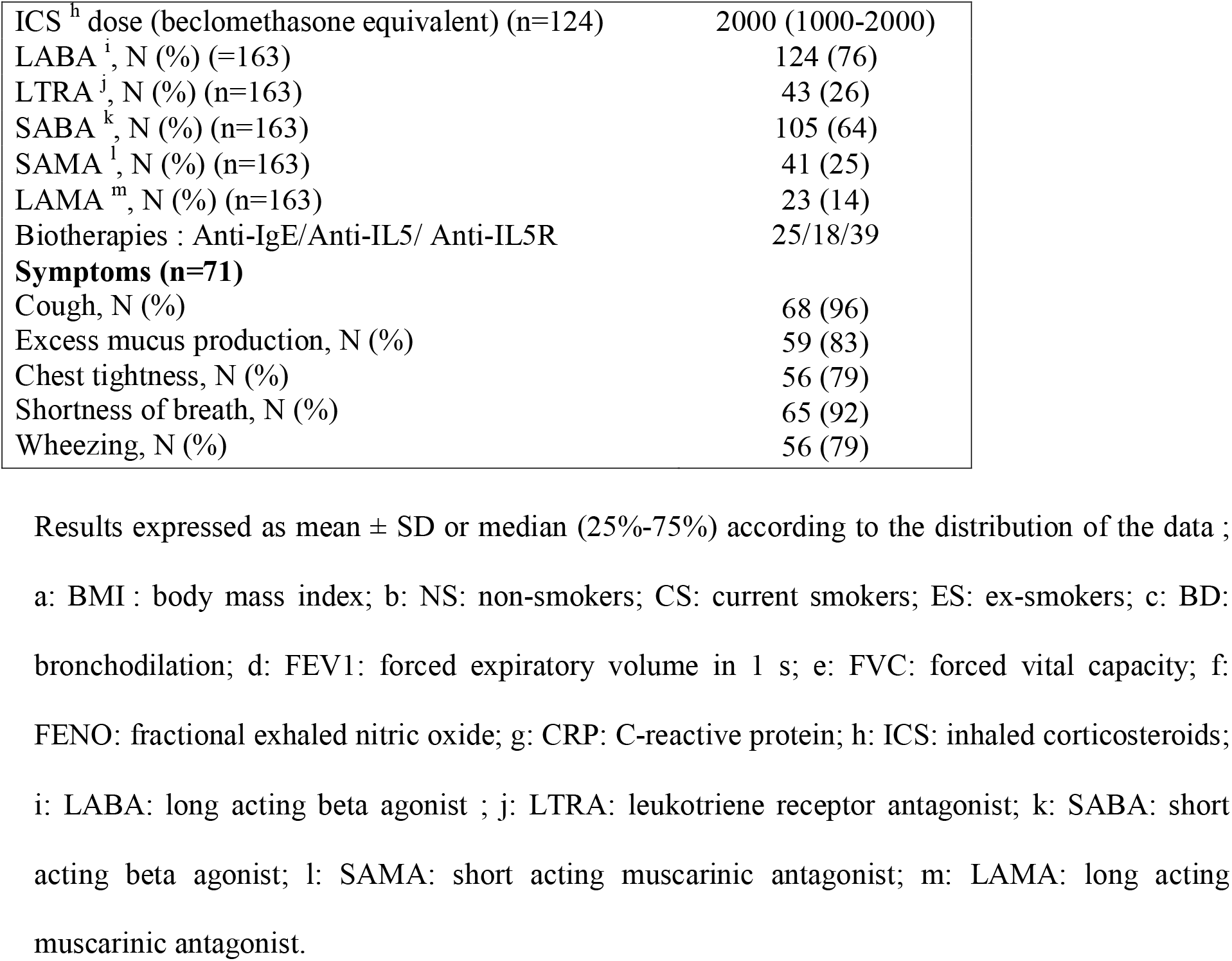
Demographic, clinical and inflammatory characteristics (n=175)

This study was approved by the Ethics committee of CHU Liege (CHU 2009/161 and 2020/107) and all subjects gave written informed consent for participation.

The samples were processed in a BSL-2 facility as previously described [9]. The sputum supernatant was obtained for all, the cell pellet containing 2. 10^6^ cells kept in RNAprotect cell reagent (Qiagen, Hilden, Germany) for 16 samples, all samples were stored at −80°C until use. We used a saliva sample collection device filled with an extraction buffer that was developed by the University of Liège and commercialized by Diagenode s.a. (Seraing, Belgium) to detect the presence of the virus in the sputum samples, supernatants and cell pellets. A previous test with serial dilution indicated that a ratio sputum/extraction buffer of 1:8 was optimal for a good detection without consuming too much sample. The method for the detection was already detailed in a recent paper [10] and is based on an inactivation of the virus and ribonucleic acid (RNA) extraction using magnetic beads followed by RT-qPCR assay that targeted the open reading frame 1ab (ORF1ab) region, the spike protein gene (S) and the nucleoprotein (N) gene of the coronavirus SARS-CoV-2. The samples were considered positive when 2 targets displayed a cycle threshold value of 37 or less. For positive control, we used a sputum from an hospitalized patient with COVID-19 pneumonia. The processing of this positive sample was made in a BSL-3 core facility.

## 3. Theory

Asthma is a common chronic airway inflammatory disease with symptoms close to those of a COVID-19 disease. In order to assess and phenotype the airway inflammation of asthmatic patients, we induced sputum even during the COVID-19 pandemic in our asthma clinic. The goal of this study was plural: to evaluate the contamination of asthmatic patients with symptoms close to COVID-19 and to find an easy way to assess the sputum samples that cannot be manipulated after processing outside a BSL-2 or BSL-3 facility. The use of a saliva sampling device is a simple detection test of interest for maintaining essential activities such as research using sputum supernatants (for mediator measurement for instance) or cells (to perform gene expression analysis among others). The loss of precious and safe samples could then be avoided.

## 4. Results

Our positive control showed a high virus load which makes us confident in the identification of positive results in the sputum samples according to our processing protocol (for the 3 genes: close to 24 cycles threshold (Ct) for the cell pellet and close to 28 Ct for the sputum supernatant). In contrast, out of all samples from the asthma clinic that were analyzed during the epidemic, none was positive for the SARS-CoV-2.

## 5. Discussions

We did not identify any positive PCR on sputum samples from asthmatic patients reporting chronic/episodic respiratory symptoms similar to what is seen in case of COVID-19 such as cough, dyspnea/shortness of breath and chest pain but without fever during the epidemic in our asthma clinical center. This indicates that if we exclude patients with body temperature > 37°C, the risk of sampling a patient with an active COVID-19 infection is extremely limited.

## 6. Conclusion

In summary, we have here described an useful technique to assess the contamination of sputum samples that have been generated during the COVID-19 period. We believe that our method of detection, due to its technical simplicity could be implemented in all centers working with induced sputum and, thereby, avoid the loss of precious samples suitable for research activities.

## Data Availability

All data produced in the present study are available upon reasonable request to the authors

## Abbreviations

ATS: American Thoracic Society
BSL: Biosafety level
COVID-19: Coronavirus infectious disease 19
Ct: Cycle threshold
ERS: European Respiratory Society
ICU: Intensive care unit
OCS: Oral corticosteroids
ORF1ab: Open reading frame 1a region
N: Nucleoprotein
RNA: Ribonucleic acid
SARS-CoV-2: Severe Acute Respiratory Syndrome Coronavirus 2
S: Spike protein gene

## Acknowledgments

The authors would like to thank Uliege/CHU Liege and the GIGA viral vector platform and Diagnostic COVID-19 platform Uliege-Cellular and molecular immunology laboratory of Uliege.

